# Development and validation of an early warning score (EWAS) for predicting clinical deterioration in patients with coronavirus disease 2019

**DOI:** 10.1101/2020.04.17.20064691

**Authors:** Yabing Guo, Yingxia Liu, Jiatao Lu, Rong Fan, Fuchun Zhang, Xueru Yin, Zhihong Liu, Qinglang Zeng, Jing Yuan, Shufang Hu, Qiongya Wang, Baolin Liao, Mingxing Huang, Sichun Yin, Xilin Zhang, Rui Xin, Zhanzhou Lin, Changzheng Hu, Boliang Zhao, Ridong He, Minfeng Liang, Zheng Zhang, Li Liu, Jian Sun, Lu Tang, Lisi Deng, Jinyu Xia, Xiaoping Tang, Lei Liu, Jinlin Hou, on behalf of Guangdong Province Working Group for COVID-19

## Abstract

**Background:** Since the pandemic outbreak of coronavirus disease 2019 (COVID-19), the health system capacity in highly endemic areas has been overwhelmed. Approaches to efficient management are urgently needed. We aimed to develop and validate a score for early prediction of clinical deterioration of COVID-19 patients.

**Methods:** In this retrospective multicenter cohort study, we included 1138 mild to moderate COVID-19 patients admitted to 33 hospitals in Guangdong Province from December 27, 2019 to March 4, 2020 (N =818; training cohort), as well as two hospitals in Hubei Province from January 21 to February 22, 2020 (N =320; validation cohort) in the analysis.

**Results:** The 14-day cumulative incidences of clinical deterioration were 7.9% and 12.1% in the training and validation cohorts, respectively. An Early WArning Score (EWAS) (ranging from 0 to 4.5), comprising of age, underlying chronic disease, neutrophil to lymphocyte ratio, C-reactive protein, and D-dimer levels, was developed (AUROC: 0.857). By applying the EWAS, patients were categorized into low-, medium-, and high risk groups (cut-off values: two and three). The 14-day cumulative incidence of clinical deterioration in the low-risk group was 1.8%, which was significantly lower than the incidence rates in the medium-(14.4%) and high-risk (40.9%) groups (P <.001). The predictability of EWAS was similar in the validation cohort (AUROC =0.781), patients in the low-, medium-, and high-risk groups had 14-day cumulative incidences of 2.6%, 10.0%, and 25.7%, respectively (P <.001).

**Conclusion:** The EWAS, which is based on five common parameters, can predict COVID-19-related clinical deterioration and may be a useful tool for a rapid triage and establishing a COVID-19 hierarchical management system that will greatly focus clinical management and medical resources to reduce mortality in highly endemic areas.

## Introduction

Coronavirus disease 2019 (COVID-19) is a respiratory tract infection caused by a new coronavirus to which there is no pre-existing immunity in humans. The continued increase in the number of cases with COVID-19 and the number of affected countries over time are of great concern. On February 28, 2020, the World Health Organization (WHO) raised the international risk assessment for the COVID-19 outbreak from “high” to “very high” nearly a month after the novel coronavirus was declared a public health emergency of international concern, which is the WHO’s highest level alarm.^1^ As of April 13, 2020, the global number of reported cases of COVID-19 has reached 1 773 084, with 83 597 cases in China and 1 689 487 cases outside of China.^2^

Hubei Province was the center of the epidemic area in the early stage of the pandemic. As of April 13, 2020, 6 7803 confirmed cases with COVID-19 were reported in Hubei Province, ^2^ which accounted for over 80% of the cases in China, and the case fatality rate in Hubei Province was five times greater than in areas outside of Hubei Province.^3^ Moreover, approximately 26% of cases developed severe disease in Hubei Province which accounted for 98% of the severe cases in China. Guangdong Province had the second largest number of confirmed cases with COVID-19 in China outside of Hubei, with 1 564 reported cases as of April 13, 2020.^2^

Based on the clinical characteristics reported by patients with COVID-19, around 80% of the patients were diagnosed with mild or moderate COVID-19 and a part of those will develop severe disease rapidly.^4,5^ The median time from onset to clinical recovery for mild cases is approximately two weeks, and it is three to six weeks for patients with severe or critical COVID-19.^4^ Previous studies found that older patients with underlying chronic illnesses, such as hypertension and diabetes, are more likely to develop severe pneumonia. ^5-10^ Moreover, it was reported that patients with severe pneumonia in comparison to patients with mild pneumonia, frequently have elevated C-reactive protein (CRP), decreased lymphocytes, and elevated D-dimer. ^5-10^ Preliminary data from the WHO suggest that the time interval from onset of symptoms to the development of severe disease is one week.^4^ Thus, efficient and timely management of patients with a high risk of developing severe COVID-19 is crucial in the face of severe resource constraints. Currently, there are no effective prediction tools for the early stratification of COVID-19 patients according to different risks of clinical deterioration. We therefore conducted this study with the aim to develop and validate an early warning score for predicting the clinical course of patients with COVID-19. We hypothesized that this score could be used as an efficient and widely applicable evaluation tool to prioritize managing patients with a high risk of developing severe to critical COVID-19 at an early stage.

## Methods

### Subjects and Data collection

This study was based on two datasets of Chinese patients with mild to moderate COVID-19 from 35 hospitals in two Chinese provinces. The inpatients from 33 hospitals in Guangdong Province were used as the training dataset to derive a score in predicting clinical deterioration of COVID-19 within 14 days after admission, whereas the inpatients from Wuhan Hankou Hospital (Wuhan, Hubei Province, China) and Honghu People’s Hospital (Jingzhou, Hubei Province, China) were used for external validation of the scoring system. All patients were diagnosed and treated according to the Chinese Guidance for COVID-19.^11^ The data, including demographic characteristics, clinical signs and symptoms, laboratory results, chest computer tomography (CT) scan images and clinical outcomes, were collected. The data of patients in Guangdong Province were extracted from the information reporting system established by the Health Commission of Guangdong Province. The data of patients in Hubei Province were extracted from the hospital information system, nursing records, and laboratory reports of the participating hospitals.

### Definitions

According to the Chinese COVID-19 prevention and control program (7^th^ edition),^11^ a confirmed case was defined by the presence of severe acute respiratory syndrome coronavirus 2 (SARS-CoV-2) in respiratory specimens detected by real-time reverse transcription polymerase chain reaction and/or positive results of immunoglobulin [Ig] M or IgG to SARS-CoV-2 testing. Mild COVID-19 was defined as having mild symptoms without radiographic evidence of pneumonia. Moderate COVID-19 was defined as having fever or respiratory symptoms with radiographic evidence of pneumonia. Severe COVID-19 was defined as meeting one of the following criteria: 1) presence of shortness of breath with a respiratory rate ≥ 30 breaths/minute; 2) an oxygen saturation (SpO_2_) ≤ 93% in the resting state; 3) hypoxemia defined as an arterial partial pressure of oxygen divided by the fraction of inspired oxygen (PaO_2_/FiO_2_ ratio) ≤ 300 mmHg; or 4) evidence of radiographic progression, defined as a ≥ 50% increase of target lesion within 24-48 hours. Critical COVID-19 was defined as meeting one of the following criteria: 1) respiratory failure plus mechanical ventilation; 2) circulatory shock; or 3) a combination of multiple organ failure plus the need for intensive care. In this study, clinical deterioration was defined as meeting the criteria of severe or critical COVID-19 in patients with mild or moderate COVID-19 at admission. The analysis time was the time interval between the date of admission and the date of clinical deterioration or the end of follow-up in the absence of clinical deterioration.

### Statistical analysis

All data were entered into and analyzed using the Statistical Package for Social Science (SPSS version 20.0, Chicago, IL, USA) and R (Version 3.5.1). Data are expressed as counts and percentages for categorical variables and as medians (interquartile ranges [IQRs]) for continuous variables. Qualitative and quantitative differences between groups were analyzed using chi-square or Fisher’s exact tests for categorical parameters and Mann-Whitney’s tests for continuous parameters, as appropriate. The cumulative probabilities of clinical deterioration were estimated by the Kaplan-Meier method and compared with the log-rank test. Univariable and multivariable Cox proportional hazards regression models were used to estimate the effect of various variables on the hazard of clinical deterioration. Hazard ratios (HR) and their 95% confidence intervals (CI) together with corresponding p values are presented. A p value of <.05 was considered to be statistically significant.

The study was conducted in accordance with the guidelines of the Declaration of Helsinki and the principles of International Committee of Harmonization (ICH) Good Clinical Practice (ICH - GCP). and was approved by the Nanfang Hospital, Guangzhou, Guangdong Province Ethics Committee (NFEC-2020-026).

## Results

### Patient Characteristics at admission

During the study period, 1043 and 662 patients were admitted to the hospitals in Guangdong and Hubei Provinces, respectively. After excluding patients with pre-existing severe to critical COVID-19 at admission (N =158, 133), patients who had incomplete medical records or laboratory tests (N = 61, 199), and patients co-infected with other respiratory viruses (N = 6, 10), the final analysis included 818 and 320 patients with mild to moderate COVID-19 from Guangdong (training cohort) and Hubei (validation cohort) Provinces, respectively (Supplementary Figure 1). Overall, 725 of 817 patients (88.7%) from Guangdong and 306 of 311 (98.4%) patients from Hubei province had images consistent with pneumonia on chest CT in the training and validation cohorts, respectively. In both cohorts, patients who experienced clinical deterioration were older, had higher neutrophil counts, lower lymphocyte counts, higher C-reactive protein levels, and higher D-dimer levels. Male patients and those with underlying chronic conditions had a higher probability of experiencing clinical deterioration. More patients with clinical deterioration received antibiotic and corticosteroid treatment (Table 1).

**Table 1.**
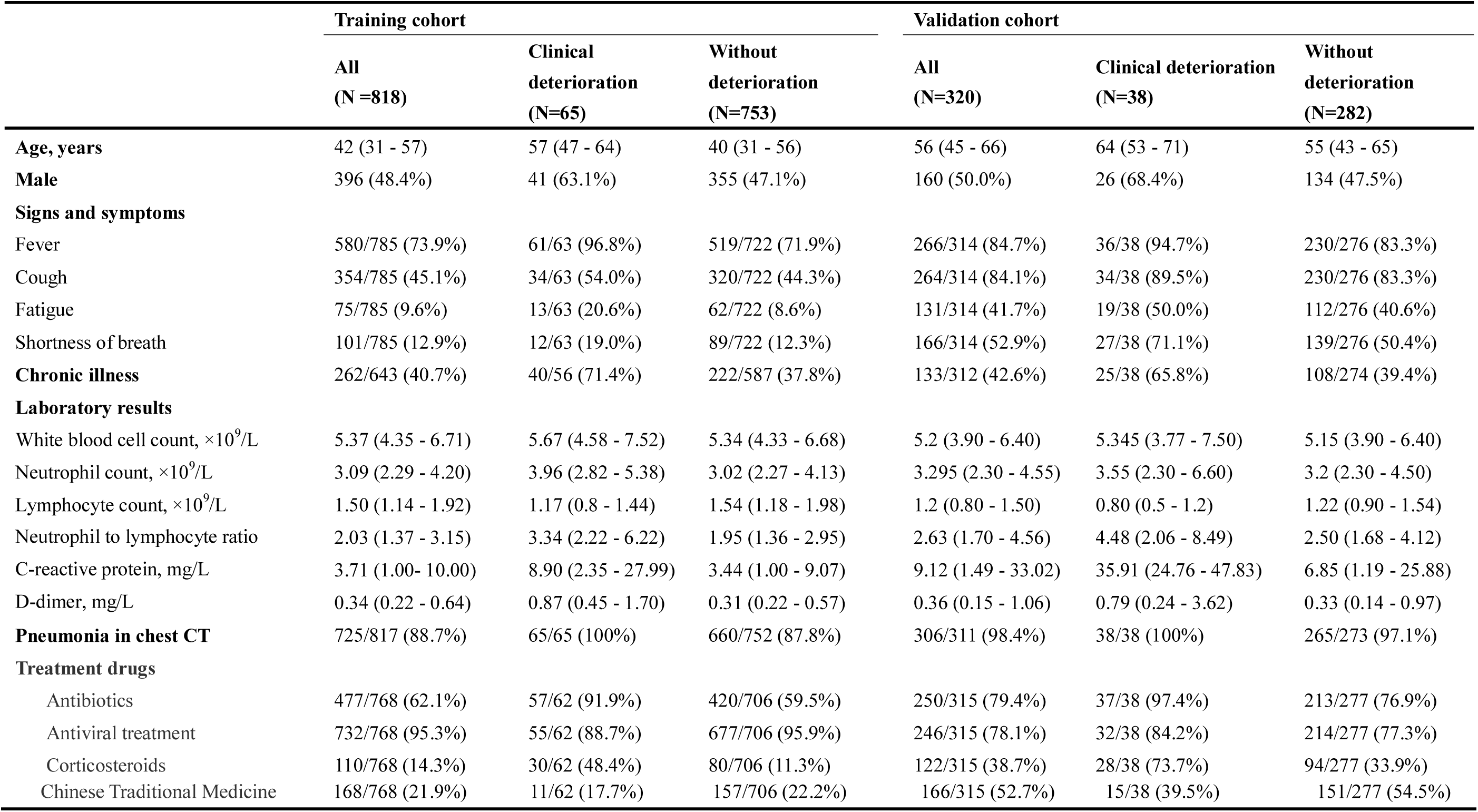
Clinical characteristics of patients with Coronavirus Disease 2019 in the training and validation cohort on admission.

|  | Training cohort |  |  | Validation cohort |  |  |
| --- | --- | --- | --- | --- | --- | --- |
|  | All<br>(N=818) | Clinical<br>deterioration<br>(N=65) | Without<br>deterioration<br>(N=753) | All<br>(N=320) | Clinical deterioration<br>(N=38) | Without<br>deterioration<br>(N=282) |
| **Age, years** | 42 (31 - 57) | 57 (47 - 64) | 40 (31 - 56) | 56 (45 - 66) | 64 (53 - 71) | 55 (43 - 65) |
| **Male** | 396 (48.4%) | 41 (63.1%) | 355 (47.1%) | 160 (50.0%) | 26 (68.4%) | 134 (47.5%) |
| **Signs and symptoms** |  |  |  |  |  |  |
| Fever | 580/785 (73.9%) | 61/63 (96.8%) | 519/722 (71.9%) | 266/314 (84.7%) | 36/38 (94.7%) | 230/276 (83.3%) |
| Cough | 354/785 (45.1%) | 34/63 (54.0%) | 320/722 (44.3%) | 264/314 (84.1%) | 34/38 (89.5%) | 230/276 (83.3%) |
| Fatigue | 75/785 (9.6%) | 13/63 (20.6%) | 62/722 (8.6%) | 131/314 (41.7%) | 19/38 (50.0%) | 112/276 (40.6%) |
| Shortness of breath | 101/785 (12.9%) | 12/63 (19.0%) | 89/722 (12.3%) | 166/314 (52.9%) | 27/38 (71.1%) | 139/276 (50.4%) |
| **Chronic illness** | 262/643 (40.7%) | 40/56 (71.4%) | 222/587 (37.8%) | 133/312 (42.6%) | 25/38 (65.8%) | 108/274 (39.4%) |
| **Laboratory results** |  |  |  |  |  |  |
| White blood cell count, $\times 10^9/L$ | 5.37 (4.35 - 6.71) | 5.67 (4.58 - 7.52) | 5.34 (4.33 - 6.68) | 5.2 (3.90 - 6.40) | 5.345 (3.77 - 7.50) | 5.15 (3.90 - 6.40) |
| Neutrophil count, $\times 10^9/L$ | 3.09 (2.29 - 4.20) | 3.96 (2.82 - 5.38) | 3.02 (2.27 - 4.13) | 3.295 (2.30 - 4.55) | 3.55 (2.30 - 6.60) | 3.2 (2.30 - 4.50) |
| Lymphocyte count, $\times 10^9/L$ | 1.50 (1.14 - 1.92) | 1.17 (0.8 - 1.44) | 1.54 (1.18 - 1.98) | 1.2 (0.80 - 1.50) | 0.80 (0.5 - 1.2) | 1.22 (0.90 - 1.54) |
| Neutrophil to lymphocyte ratio | 2.03 (1.37 - 3.15) | 3.34 (2.22 - 6.22) | 1.95 (1.36 - 2.95) | 2.63 (1.70 - 4.56) | 4.48 (2.06 - 8.49) | 2.50 (1.68 - 4.12) |
| C-reactive protein, mg/L | 3.71 (1.00- 10.00) | 8.90 (2.35 - 27.99) | 3.44 (1.00 - 9.07) | 9.12 (1.49 - 33.02) | 35.91 (24.76 - 47.83) | 6.85 (1.19 - 25.88) |
| D-dimer, mg/L | 0.34 (0.22 - 0.64) | 0.87 (0.45 - 1.70) | 0.31 (0.22 - 0.57) | 0.36 (0.15 - 1.06) | 0.79 (0.24 - 3.62) | 0.33 (0.14 - 0.97) |
| **Pneumonia in chest CT** | 725/817 (88.7%) | 65/65 (100%) | 660/752 (87.8%) | 306/311 (98.4%) | 38/38 (100%) | 265/273 (97.1%) |
| **Treatment drugs** |  |  |  |  |  |  |
| Antibiotics | 477/768 (62.1%) | 57/62 (91.9%) | 420/706 (59.5%) | 250/315 (79.4%) | 37/38 (97.4%) | 213/277 (76.9%) |
| Antiviral treatment | 732/768 (95.3%) | 55/62 (88.7%) | 677/706 (95.9%) | 246/315 (78.1%) | 32/38 (84.2%) | 214/277 (77.3%) |
| Corticosteroids | 110/768 (14.3%) | 30/62 (48.4%) | 80/706 (11.3%) | 122/315 (38.7%) | 28/38 (73.7%) | 94/277 (33.9%) |
| Chinese Traditional Medicine | 168/768 (21.9%) | 11/62 (17.7%) | 157/706 (22.2%) | 166/315 (52.7%) | 15/38 (39.5%) | 151/277 (54.5%) |

### Risk factors for clinical deterioration

At a median follow-up of 18 days (IQR, 13 - 24 days), 24 patients experienced clinical deterioration with 14-day cumulative incidence of 7.9% (95% CI, 6.0% −9.7%) in the training cohort. In the univariable Cox regression analysis, sex, age, underlying chronic illness, days from onset of symptoms to admission, white blood cell count, neutrophil count, lymphocyte count, neutrophil to lymphocyte ratio (NLR), CRP and D-dimer were associated with the 14-day incidence of clinical deterioration. By multivariable Cox regression analysis (with stepwise selection), it was found that age (> 50 vs. ≤ 50 years) (hazard ratio [HR], 2.158; 95% CI, 1.136 - 4.097; *P* =.019), underlying chronic disease (Yes vs. No) (HR, 2.631; 95% CI, 1.399 - 4.951; *P* =.003), neutrophil to lymphocyte ratio (> 5 vs. ≤ 5) (HR, 2.176; 95% CI, 1.128 - 4.199; *P* =.021), C-reactive protein (> 25 vs. ≤ 25 mg/L) (HR, 2.577; 95% CI, 1.358 - 4.889; *P* =.004), and D-dimer (> 0.8 vs.≤ 0.8 mg/L) (HR, 4.567; 95% CI, 2.605 - 8.008; *P* <.001) were independently significant factors in predicting the 14-day risk of clinical deterioration (Table 2).

**Table 2.** Association between clinical characteristics and laboratory parameters and 14-day clinical deterioration in the training cohort.

| Variables | Univariable analysis |  |  | Multivariable analysis |  |  |
| --- | --- | --- | --- | --- | --- | --- |
|  | HR | 95% CI | P | HR | 95% CI | P |
| Sex (male vs. female) | 1.841 | 1.115 to 3.038 | .018 |  |  |  |
| Age (> 50 vs. ≤ 50 years) | 3.600 | 2.159 to 6.003 | <.001 | 2.158 | 1.136 to 4.097 | .019 |
| Chronic illness (Yes vs. No) | 3.838 | 2.155 to 6.833 | <.001 | 2.631 | 1.399 to 4.951 | .003 |
| Days from illness to admission (> 7 vs. ≤ 7 days) | 2.222 | 1.281 to 3.852 | .005 |  |  |  |
| White blood cell count (> 7.5 vs. ≤ 7.5 ×10<sup>9</sup>/L) | 1.833 | 1.045 to 3.213 | .036 |  |  |  |
| Neutrophil count (> 4.5 vs. ≤ 4.5×10<sup>9</sup>/L) | 2.746 | 1.672 to 4.511 | <.001 |  |  |  |
| Lymphocyte count (≤ 0.9 vs. > 0.9×10<sup>9</sup>/L) | 3.635 | 2.167 to 6.097 | <.001 |  |  |  |
| Neutrophil to lymphocyte ratio (> 5 vs. ≤ 5) | 4.308 | 2.526 to 7.346 | <.001 | 2.176 | 1.128 to 4.199 | .021 |
| C-reactive protein (> 25 vs. ≤ 25 mg/L) | 4.578 | 2.684 to 7.807 | <.001 | 2.577 | 1.358 to 4.889 | .004 |
| D-dimer (> 0.8 vs. ≤ 0.8 mg/L) | 5.687 | 3.473 to 9.314 | <.001 | 4.567 | 2.605 to 8.008 | <.001 |

**Note:** The threshold value of each continuous variable was the value with a maximum sum of sensitivity and specificity for predicting clinical deterioration.

### Refining the early warning score

An Early WArning Score (EWAS) was developed using the above identified five significant variables, for which the weights were defined as the quotient (rounded to nearest integer) of the corresponding estimated coefficient from a multivariable Cox regression analysis divided by the smallest χ^2^ coefficient (Table 3). EWAS ranged from 0 to 4.5. The AUROC was 0.857 (95% CI, 0.807 - 0.907, *p* <.001) in the training cohort. Using two and three as the cutoff values, among the 552 patients with available scores, 338 (61.2%), 146 (26.4%) and 68 (12.3%) patients were in the low-, medium-, and high-risk categories. The 14-day cumulative incidence rates of clinical deterioration were 1.8%, 14.4%, and 40.9% in the three risk groups, respectively (*p* <.001). (Figure 1A) The cut-off value of two was associated with 70.8% sensitivity and 96.9% negative predictive value (NPV). The cut-off value of three resulted in 94.6% specificity and 39.7% positive predictive value (PPV) (Table 4). The calibration plot for the 14-day probability of remaining free clinical deterioration was predicted well in the training cohort (Figure 1C).

**Figure 1.**
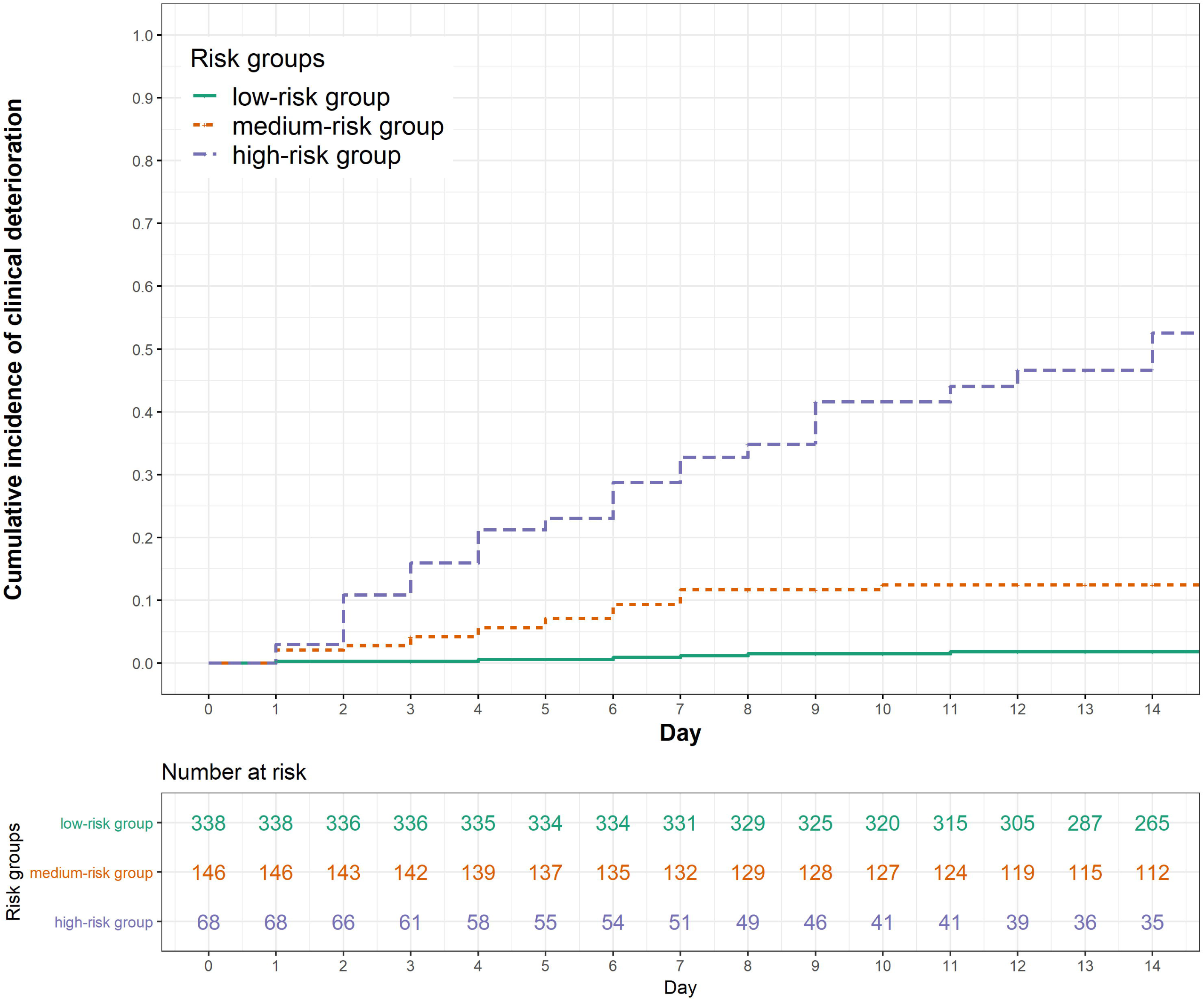
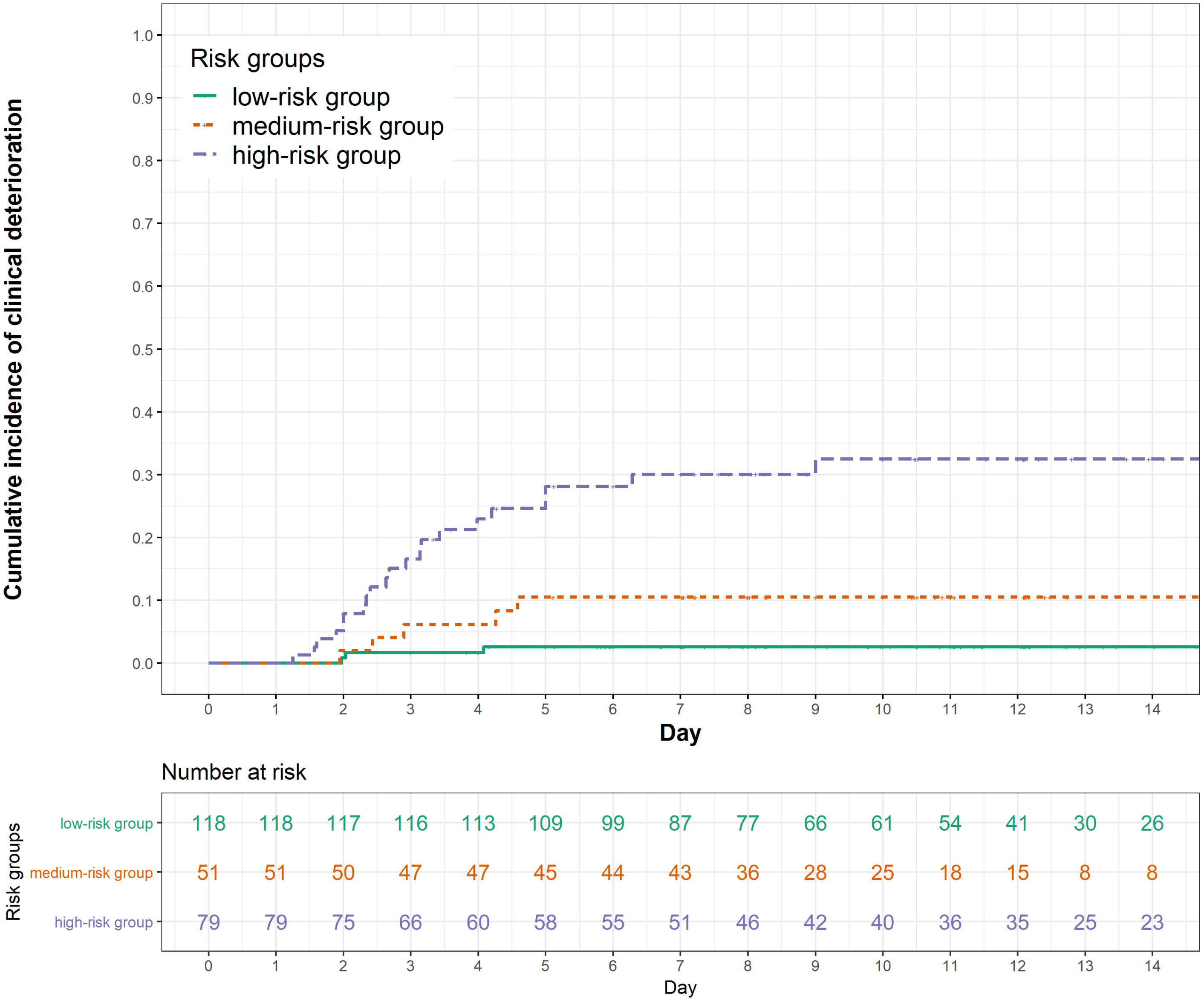
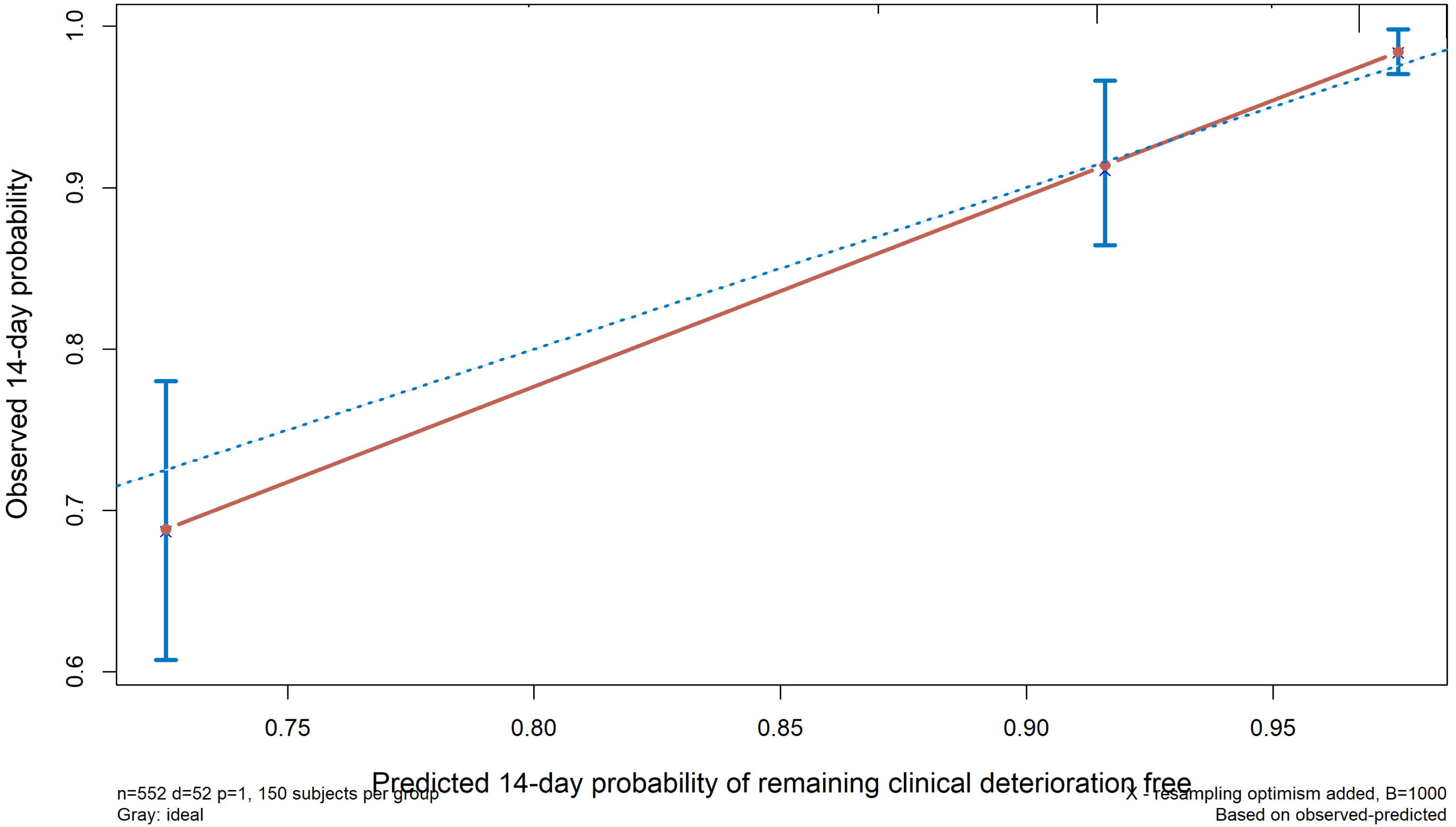
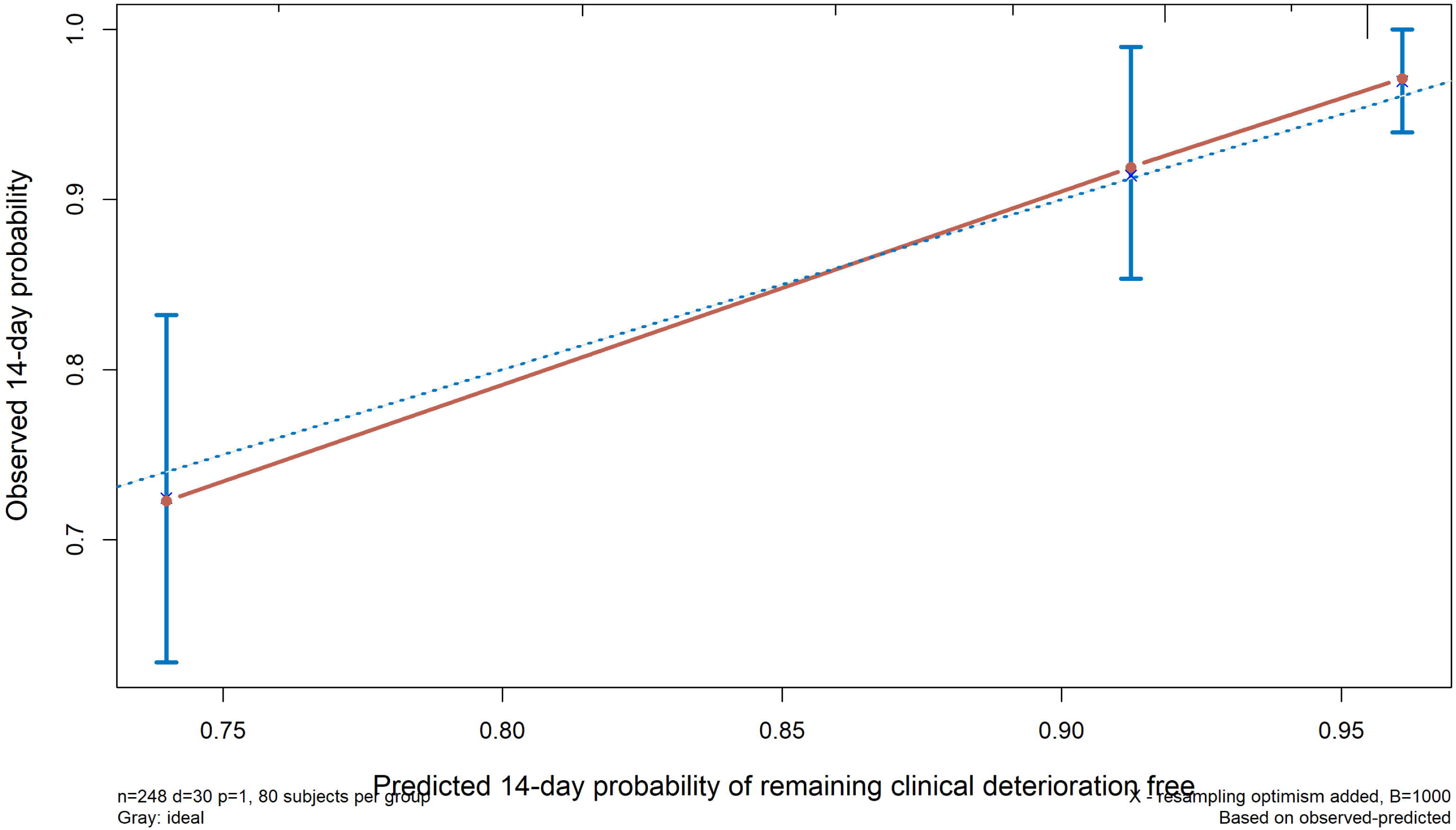
The discrimination (A, B) and calibration (C, D) curves of early warning score (EWAS) to predict clinical deterioration in each cohort.

**Table 3.** Construction of the early warning score (EWAS) for prediction of clinical deterioration in patients with COVID-19.

|  | Score |
| --- | --- |
| **Age, years** |  |
| ≤50 | 0 |
| >50 | +1 |
| **Chronic illness** |  |
| No | 0 |
| Yes | +1 |
| **Neutrophil to lymphocyte ratio** |  |
| ≤5 | 0 |
| >5 | +1 |
| **C-reactive protein, mg/L** |  |
| ≤25 | 0 |
| >25 | +1 |
| **D-dimer, mg/L** |  |
| ≤0.8 | 0 |
| >0.8 | +1.5 |

**Note:** The score was ranged from 0 to 4.5.

**Table 4.** Accuracy for prediction of clinical deterioration in the training and validation cohorts using the cut-off of two and three in the early warning score (EWAS).

|  | Training cohort |  |  |  | Validation cohort |  |  |  |
| --- | --- | --- | --- | --- | --- | --- | --- | --- |
|  | Cut-off value: 2 |  | Cut-off value: 3 |  | Cut-off value: 2 |  | Cut-off value: 3 |  |
|  | Value | 95% CI | Value | 95% CI | Value | 95% CI | Value | 95% CI |
| Sensitivity, % | 70.8 | 59.7, 81.8 | 41.5 | 29.6, 53.5 | 71.1 | 56.6, 85.5 | 57.9 | 42.2, 73.6 |
| Specificity, % | 77.7 | 74.7, 80.7 | 94.6 | 92.9, 96.2 | 63.5 | 57.9, 69.1 | 79.8 | 75.1, 84.5 |
| PPV, % | 21.5 | 16.0, 27.0 | 39.7 | 28.1, 51.3 | 20.8 | 13.8, 27.7 | 27.8 | 18.0, 37.7 |
| NPV, % | 96.9 | 95.5, 98.2 | 94.9 | 93.4, 96.5 | 94.2 | 90.9, 97.5 | 93.4 | 90.2, 96.5 |
| PLR | 3.2 | 2.6, 3.9 | 7.6 | 5.0, 11.5 | 1.9 | 1.5, 2.5 | 2.9 | 2.0, 4.1 |
| NLR | 0.4 | 0.3, 0.6 | 0.6 | 0.5, 0.8 | 0.5 | 0.3, 0.8 | 0.5 | 0.4, 0.8 |

### External validation of the early warning score

A total of 38 patients experienced clinical deterioration with a 14-day cumulative incidence of 12.1% (95% CI, 8.4% - 15.8%) during a median follow-up of ten days (IQR, 7 - 14 days) in the validation cohort. The AUROC of EWAS in the validation cohort was 0.781 (95% CI, 0.695 - 0.866, *p* <.001). Among the 248 patients with evaluable scores in the validation cohort, 118 (47.6%), 51 (20.6%) and 79 (31.9%) of the patients were assigned to the low-, medium-, and high-risk groups, respectively. The K-M curves also showed equally good discrimination among the three risk groups in the validation cohort. The 14-day cumulative incidences of clinical deterioration were 2.6%, 10.0%, and 25.7% in the low-, medium- and high-risk groups, respectively (*P* <.001) (Figure 1B). The cut-off value of two was associated with 71.1% sensitivity and 94.2% NPV. The cut-off value of three resulted in 79.8% specificity and 27.8% PPV (Table 4). The calibration plot of the model in the validation cohort is depicted in Figure 1D.

## Discussion

With the spread of the epidemic, an increasing number of studies describing the clinical characteristics of COVID-19 have been reported. However, there is still lack of knowledge on many aspects of COVID-19, such as clinical predictors manifestations of asymptomatic and mild cases who progress. While most people with COVID-19 develop only mild or uncomplicated illness, approximately 14%, even among the young patients, develop severe disease.^12^ Thus, it is critical to better understand the clinical features of COVID-19 and the risk of disease progression. To our knowledge, we describe here the first validated score for predicting clinical deterioration of COVID-19 which was developed with the dataset of COVID-19 patients from Guangdong Province and validated with the dataset of COVID-19 patients from Hubei Province. Our findings show that the early-warning score can be useful in the assessment of the 14-day risk of clinical deterioration in patients with mild to moderate COVID-19 at admission. A major advantage of this score is that it is practical and easy to use in routine clinical practice, based on patient’s age, history of chronic disease, NLR, CRP, and D-dimer. It can therefore be widely applicable in many health care settings globally. Although there were several differences in the characteristics of the patients in the training and validation datasets, this strengthens the reliability of our score, which was shown to offer similar predictability in different patient populations.

Age has been reported as an important independent predictor of disease progression in Severe Acute Respiratory Syndrome (SARS) and Middle East respiratory syndrome (MERS).^13,14^ Previous studies on COVID-19 also revealed that the elderly are prone to have higher incidences of severe illness and mortality. Our study confirms that increased age is also associated with disease progression in patients with mild or moderate COVID-19. Presence of underlying chronic illness is a common risk factor for progression in most diseases. The most common chronic diseases in our cohort were hypertension, pre-existing cardiac conditions, chronic lung disease and diabetes, which had prevalence rates of 12.6%, 12.2%, 7.3% and 5.5% in the training cohort, respectively. These comorbidities are found to be important risk factors in previously reported seriously ill or non-survival COVID-19 patients. In addition, patients with these illnesses may receive angiotensin-converting enzyme (ACE) inhibitor treatments. These regimens increase the expression of ACE2, which is the receptor of SARS-CoV-2 and may contribute to disease progression.^15,16^

One of the reported early general manifestations in the course of SARS-CoV-2 infection is lymphocytopenia in the presence of a normal WBC count and more prominent abnormalities in lymphocyte counts are identified in patients with more severe pneumonia. In the present study, a correlation between a higher neutrophil count and early deterioration was also observed. Hence, we included the NLR in the multivariable regression analysis. Additionally, NLR is used as a predictor of community acquired pneumonia (CAP), specifically in elderly patients.^17-19^ CRP is a common indicator of the inflammatory response. Its prediction value of disease progression has been reported in MERS, influenza-infected and CAP patients. ^20-22^ In our study, a CRP level higher than 25 mg/L at admission may indicate an underlying secondary infection or an intense inflammatory response that may lead to disease progression in the short term. Another factor, D-dimer, is associated with coagulation activation, which has been previously been reported to be a common finding in COVID-19 patients who had severe illness or died. Additionally, elevated D-dimer is one of the risk factors for the development of acute respiratory distress syndrome (ARDS), progression from ARDS to death or early in-hospital death in COVID-19 patients. ^23,24^ The possible mechanisms include ischemia and thrombosis caused by systemic proinflammatory cytokine responses, which are reported to be mediators of atherosclerosis directly contributing to plaque rupture through local inflammation. ^25,26^

Management decisions for the high-risk patients with COVID-19 must be made quickly and are often based upon scant evidence. Under these pressing clinical circumstances, high quality and timely evidence-based guidance becomes especially crucial. EWAS, with a high level of differentiating power, can be regarded as such quality evidence in providing an efficient, feasible and practical approach to establish a hierarchical management system of COVID-19 in highly endemic SARS-CoV-2 areas where medical resources are extremely limited. Specifically, it is suggested that, by applying the EWAS, patients classified in the low-risk group should be isolated and treated in “mobile cabin hospitals”, an isolation facility converted from sports stadiums, convention centers, etc. with structures easily installed which can accommodate large numbers of patients. Patients in the medium-risk group should be treated in the general wards, whereas patients in the high-risk group should be transferred to the intensive care unit for more comprehensive treatment due to the high risk of clinical deterioration within 14 days of admission being as high as approximately 40% in this group. It is predicted that this approach would benefit a considerable number of patients with COVID-19 by directing the appropriate level of medical resources according to the severity of the disease, thus reducing mortality and saving valuable medical and socioeconomic resources.

Our study has several strengths, including the use of two independent cohorts with large sample sizes, which increase the reliability of the results. Nonetheless, the study also has certain limitations. First, the patients in the training cohort were enrolled from more than 30 hospitals in Guangdong Province, and their disease status, exposure history, and treatment strategy were relatively heterogeneous; however, this heterogeneity strengthens the reliability of our scoring methodology, which shows similar predictive ability in different patient populations. Secondly, the EWAS was based on artificially defined categorical variables, which may have led to the loss of detailed continuous data. However, the categorical variables will be much simpler to apply and promote in highly endemic areas. Thirdly, the patients in the two cohorts were all Chinese. Whether the study results are applicable to patients in other Eastern or Western countries merits further investigation.

In conclusion, the early-warning score, which is based on patients’ age, underlying chronic disease, NLR, CRP, and D-dimer, represents a reliable and simple scoring system for the prediction of clinical deterioration of COVID-19 within 14 days after admission. It may be a useful and convenient tool for a rapid triage and establishing a hierarchical management system of COVID-19 patients that will greatly focus clinical management and medical resources to reduce mortality in highly endemic areas.

## Data Availability

All data included were available.

## Author’s Contribution

Concept and design: Jinlin Hou, Yabing Guo

Data collection: Yingxia Liu, Jiatao Lu, Shufang Hu, Fuchun Zhang, Qinglang Zeng, Jing Yuan, Qiongya Wang, Baolin Liao, Mingxing Huang, Sichun Yin, Xilin Zhang, Rui Xin, Zhanzhou Lin, Changzheng Hu, Boliang Zhao, Ridong He, Mingfeng Liang, Lisi Deng, Jinyu Xia

Data analysis: Rong Fan, Zhihong Liu, Xueru Yin

Drafting of the manuscript: Rong Fan, Zhihong Liu, Xueru Yin

Critical revision of the manuscript: Jian Sun, Li Liu, Lu Tang

Supervision: Jinlin Hou, Lei Liu, Xiaoping Tang

## Acknowledgement

We thank all the staff of participating hospitals and pay tribute to all the medical staff who are currently on the front line of the epidemic. We thank George F Gao for his critical revision and professional advice on this manuscript.

